# Built environment’s impact on COVID-19 transmission and mental health revealed by COVID-19 Participant Experience data from the All of Us Research Program

**DOI:** 10.1101/2022.04.05.22273358

**Authors:** Wenting Luo, Edwin Baldwin, Anna Yi Jiang, Shujuan Li, Bo Yang, Haiquan Li

## Abstract

**Objectives:** The coronavirus disease 2019 (COVID-19) pandemic has led to millions of deaths. Effectively cutting the transmission of COVID-19 is essential to reduce the impact. Previous studies have observed the potential relationship between the built environment and COVID-19 transmission; however, to date, stringent studies investigating these relationships at the individual level are still insufficient. Here, we aim to examine the relationship between household types and COVID-19 infection (or mental health) during the early stages of the pandemic by using the *All of Us* Research Program **CO**VID-19 **P**articipant **E**xperience (COPE) survey data.

**Design:** Based on 62,664 participants’ responses to COPE from May to July 2020, we matched the cases of self-reported COVID-19 status, anxiety, or stress, with controls of the same race, sex, age group, and survey version. We conducted multiple logistic regressions between one of the outcomes and household type under the adjustment of other related covariates, such as ethnicity, age, social distancing behavior, and house occupancy.

**Results:** Household type with a shared component was significantly associated with COVID-19 infection (OR=1.19, 95% CI 1.1 to 1.3; p=2×10^−4^), anxiety (OR=1.26, 95% CI 1.1 to 1.4; p=1.1×10^−6^), and stress (OR=1.29, 95% CI 1.2 to 1.4, p=4.3×10^−10^) as compared to free-standing houses after adjusting for the abovementioned confounding factors. Further, frequent nonessential shopping or outings, another indicator of the built environment, was also associated with COVID-19 infection (OR=1.36, 95% CI 1.1 to 1.8; p=0.02), but not associated with elevated mental health conditions.

**Conclusion:** Our study demonstrated that the built environment of houses with a shared component tends to increase the risk of COVID-19 transmission, which consequently led to more anxiety and stress for their dwellers. It also suggested the necessity to improve the quality of the built environment through planning, design, and management toward a more resilient society in coping with future pandemics.

## 1 Introduction

Since the start of the COVID-19 pandemic in December of 2019, it has resulted in more than six million reported deaths. The virus of SARS-CoV-2 is transmitted via inhalation of the virus in air that has been contaminated with the respiratory fluids of infected persons which are released as particles and droplets^1^. It can also be circulated in aerosols^2^ if the air ventilation rate is insufficient^3^ or if the air is highly recycled in a closed setting, such as a plane or a cruise ship, as seen on the Diamond Princess Cruise Ship^4^. On the other hand, the virus can remain on the surface of objects, such as doorknobs, stairs, and elevator panel buttons, for hours to days^5^. Thus, the virus can be transmitted by soiled hands which have been contaminated by touching the surface of objects and then by touching the mucous membranes of their bodies (e.g., noses). In both cases, SARS-CoV-2 is transmitted within the built environment, which makes the impact of the built environment on COVID-19 transmission a critical issue. This is particularly true during the initial phase of a pandemic when the knowledge of the transmission approaches for the public is very limited.

A handful of studies have reported the connection between various types of built environments and COVID-19 transmission, as summarized in a review study^6^. These studies spanned different types of built environments from multiple countries in a variety of cities, for instance, trains in and between cities in Hubei Province (e.g., Wuhan)^7^, restaurants and public markets in Hong Kong^8,9^, transportation infrastructure in Huangzhou^10^, apartment air ducts in Seoul^11^, house quality and crowding in Washington D.C.^12^, building values, units, and membership in New York City^13^, and intervention of the built environment in cities of Turkey^14^. These studies found that housing quality and living conditions were strong predictors for the ward level COVID-19 death count, such as in Washington D.C.^12^. Other studies have corroborated the results. For instance, a study in King County, Washington, demonstrated that built environment density was positively associated with COVID-19 incidence rates^15^. Indeed, the mitigation of viral transmission through the air delivery system can reduce the transmission of the virus. Nevertheless, current studies on the relationships between the built environment and COVID-19 transmission are still plagued by small sample sizes^13^, lack of precision, and relying on reports at county^16^, city^7^, ZIP code^15^, and community levels^12^. Stringent studies at the individual level are still lacking, most likely due to the high cost of acquiring data and the difficulty of controlling confounding covariates in prior methods.

Recent advances in data-driven projects such as the All of US Research Program (AllofUsRP)^17^, the largest biobank project in the United States, have provided unique opportunities to investigate the impact of the built environment on COVID-19 transmission. AllofUsRP has conducted six versions of large-scale, comprehensive survey studies for the COVID-19 Participant Experience (COPE) in 2020 and 2021^18^. The first three versions of COPE collected household type information and were conducted in May, June, and July of 2020, the starting period of the pandemic. These versions involved 62,664 participants with individual-level household building type information, ranging from free-standing houses to various types of apartments and studios. More importantly, COPE also tracked the mental health of the participants, plus their social distancing behaviors during 2020-2021, providing further opportunities to examine the impact of the built environment on the stress that dwellers experienced. Thus, in the current investigation, we perform a stringent association study between the built environment and COVID-19 transmission, plus its impact on residents’ mental health^19^. Higher levels of stress and anxiety have been reported due to COVID-19^20^; thus, the relevance to household type is worth investigating. We will control various types of confounders while studying the interwoven factors, taking advantage of the rich information collected by the survey. Unveiling these relationships will not only demonstrate the importance of mitigation strategies for COVID-19 transmission but will also provide adaptive and resilient design solutions focusing on the built environment to respond to potential airborne and contagious viruses in the future^21^.

As follows, we will briefly discuss the methodology of the study in Section 2, followed by detailed results in Section 3. We will then discuss the results and relationships to related studies and conclude at the end.

## 2 Methods

### 2.1 COPE Survey data in All of Us Research Program and Preprocessing

We chose the AllofUsRP dataset because of its large cohort, diversity, and availability of household type data in the COPE survey. AllofUsRP aims to recruit adults (18 years and older) who live in the United States from all backgrounds. To date, the project has enrolled over 300,000 participants, which are from diversified backgrounds (e.g., ethnicities, social behaviors, geographic locations, medical conditions) and represent their community in research studies^22^. The transparency, diversity, and inclusion of the AllofUsRP provide researchers with a unique opportunity to investigate the potential roles that the built environment might play on mental and physical health^23^. AllOfUsRP shares the data assets collected from the participants in a common cloud environment, the All of Us Researcher Workbench^22^. Qualified researchers have access to the dataset and can retrieve and analyze the data through web-based tools and interactive cloud-based computing environments^17^. Access to de-identified individual-level data requires registration and training, which has been fulfilled by all authors directly analyzing the data. The Institutional Review Board of the University of Arizona waived ethical approval for this work due to using de-identified data.

The AllofUsRP COVID-19 Participant Experience survey is an online survey that began in May 2020 and ended in February 2021, with the aim to better understand how COVID-19 affects participants’ daily lives and health conditions, especially their mental health^24^. This survey is about 20- to 30-minutes long and covers topics about social distancing experiences, self-reported COVID-19 status, well-being, basic participant’s information, mental health, COVID-19 induced socioeconomic changes (e.g., work and financial changes), and physical activity, among many others^18^. The survey had six versions and had asked the participants to answer the most recent version of questions, enabling researchers to examine the changing effects of COVID-19 over time. At the time of our analysis, we took advantage of the first three versions (May, June, and July 2020) of the COPE data consisting of 62,664 participants in AllofUsRP Dataset v4 where household type information was available. Our analysis focuses on the responses of these participants about the topics of anxiety, stress, house types, social distancing behaviors, and their COVID-19 infection conditions. We used Structural Query Language (SQL) to extract the data guided by the concept ids of the variables (outcomes and covariates; details shown below) of our interest.

### 2.2 Construction of Study Cohort

Based on participants’ responses to COPE, we conducted a retrospective case-control study and started by identifying all positives and negatives for each outcome, specifically COVID-19, anxiety, and stress status. COVID-19 status was self-reported by the question ‘Do you think you have had COVID-19’. The status was considered as the binary outcome variable (‘Yes’: positive; ‘No’: negative); we removed the participants who were not sure about their COVID-19 status and reported ‘Maybe’ for convenience. We treated ‘more than half of days’ and ‘nearly every day’ as positive anxiety cases, and ‘not at all’ as negative controls for whether participants felt nervous, anxious, or on edge during the last two weeks. We removed missing data and the response of ‘several days’ due to not being distinctly positive or negative. Similarly, we treated ‘fairly often’ and ‘very often’ as positive stress cases versus ‘never’ and ‘almost never’ as negative controls in the question of feeling nervous and stressed in the last month, excluding ‘sometimes’ from the analysis.

We matched each positive outcome cohort with a control cohort, with the same race, sex, age group (per 10 years), and survey version, similar to our previous COVID-19 study^25^. This is because these factors are related to social behaviors that directly underline COVID-19 transmission. We divided the available cases and controls of an outcome (e.g., COVID-19 infections) in COPE into strata of the same race, sex, age group, and survey version. Then, we randomly selected the same number of control individuals without replacement as the number of cases from the same stratum. In a few scenarios without enough controls, we loosened the matched field in the order of survey version, race, sex, and age group. Cases in the strata without any matched controls (including partial matches) were excluded from the analysis. Of note, if the confounding factors are fully matched, they will not be expected to be significant in the model even if they are significant before matching.

### 2.3 Statistical Analysis

We fitted the multiple logistic regression model using the COPE survey to determine the relationship between household types and one of the influenced outcomes, COVID-19 infection, anxiety, or stress during the early stages of the COVID-19 pandemic. Three models were tested with potential confounders to adjust the association explained by the covariates that we focused on. 1) Model A used in Section 3.1 analyzed COVID-19 status as the outcome given household type as the major explanatory variable (covariate), which was adjusted for other confounding covariates such as household occupancy, race/ethnicity, sex, age, social distancing, hygiene behaviors (e.g., shopping/outings), and mental health (anxiety and stress). 2) Model B and C in Section 3.2 used mental health (anxiety and stress) as the outcome to study the contribution from household types, which is adjusted for the other covariates, similar to Model A.

For household type, we examined individual types (e.g., free-standing houses and two-bedroom apartments), in addition to combining those with a shared component for a summarized analysis. This specifically included the townhouse, three- (or more), two-, and one-bedroom apartments, studio, and nursing home or rehab facilities. We also included the number of occupants in a house as a confounding covariate. For social distancing behavior, we included the number of days with the following behaviors in the last five days: staying at home, working or volunteering outside the home, attending social gatherings outside of more than 10 people, and having close contact with somebody in a risk group. We also included hygiene practices in the analysis. We treated each variable as numeric (e.g., age) or categorical data, particularly for those with missing values (missing value as a special level). The details of the studied questions and multichoice response options can be referred to on the AllofUsRP website^24^.

Odds ratios of the outcome by the major covariates were calculated from the contrasts that used conventionally selected reference groups (i.e., the free-standing house for house types; none of the days (0 days) of just for fun shopping and outings) for nonessential behavior in all models. The covariates that were irrelevant after ANOVA analysis (p>0.25 with Chi-square test) were excluded from the final logistic regression model for further analysis. All statistical tests were 2-sided, and a p-value < 0.05 was considered statistically significant. Analyses were implemented in the All of Us Researcher Workbench with R.

## 3 Results

### 3.1 Associations between household type and COVID-19 status

We first investigated whether household type (e.g., apartment) had an impact on COVID-19 transmission using the matched cases and controls (Methods 2.2). Of the 62,664 participants in the COPE Survey, 4,870 COVID-19 cases were reported from 3,700 participants (some participants reported multiple times in different versions of the survey during May, June, and July 2020). We thus matched 4,870 negative controls with the same race, sex, age group (per 10 years), and survey version (month) as the cases (four failed to match survey version and nine failed to match survey version and race). The majority of participants lived in the free-standing house (single house): 59.3% (2,886 out of 4,870) of the self-claimed cases (Covid = 1), which is close to the national average. The characteristics of the cases and matched controls are summarized in Table 1. Since free-standing houses are assumed to be less risky, we used it as the reference level to compute the risk for participants living in other house types. Using a multiple logistic regression model, we examined the association between household type and COVID-19 status, with additional controls for ethnicity, birth year, social distancing behavior, and household occupancy, plus anxiety and stress status.

**Table 1.** Characteristics of COVID-19 matched cohort from AllofUsRP COPE data collected in May, June, and July 2020

| Characteristic | COVID-19 Status, No. |  |
| --- | --- | --- |
|  | Positive | Negative |
| Age |  |  |
| 18-29 | 480 | 476 |
| 30-39 | 851 | 847 |
| 40-49 | 835 | 837 |
| 50-59 | 1113 | 1115 |
| 60 and over | 1591 | 1595 |
| Sex at birth |  |  |
| Female | 3459 | 3465 |
| Male | 1382 | 1381 |
| Others | 29 | 24 |
| Race and ethnicity |  |  |
| White | 3833 | 3843 |
| Asian | 105 | 102 |
| Black/African American | 316 | 316 |
| More than one population | 112 | 112 |
| Others* | 466 | 465 |
| Skip, prefer not to say, or no answer | 38 | 32 |

\*Including another single population, none of these, none indicated, and no matching concept

The distribution of house types conditioned on the COVID-19 status is depicted in Fig. 1, showing the probability of each household type given a COVID-19 status. The figure demonstrated that participants who became infected had a lower probability of living in a free-standing house as compared to the non-infected. Participants with positive cases were more likely to live in household types with shared components (See the reverse trend in Figure 1 for apartments). A multiple logistic regression further confirmed the trend. Participants living in household types with a shared component (e.g., townhouse and apartment) had a statistically significant higher risk of infection (OR=1.19 95% CI 1.1 to 1.3; p=0.0002) as compared to those living in a free-standing house. Among all the house types with a shared component, the odds of contracting COVID-19 for participants who live in a nursing home or rehab facility were more than seven-fold greater than participants who live in a free-standing house (OR = 7.13 95% CI 1.5 to 33.7; p = 0.01). Other significantly higher odds appear among the respondents who live in three-bedroom (or more) apartments (OR = 1.37, 95% CI 1.1 to 1.7, p= 0.001). This effect was not confounded by age which had a different infection risk, likely due to distinct social behaviors. Even when controlled by age group (using both cases and controls within the same age group), the trends of odds ratios were similar (data not shown). Some household types lacked significance, such as those depleted due to age groups (e.g., nursing home facilities for ages 40 and below). Besides, race, sex, age, ethnicity, survey version, and household occupancy were not associated with COVID-19 infection in the matched cohort, but social distancing behavior (e.g., number of days at home, gatherings with over 10 people, shopping, etc.), and mental health (anxiety and stress) were (discussed in the next sections).

**Fig. 1.**
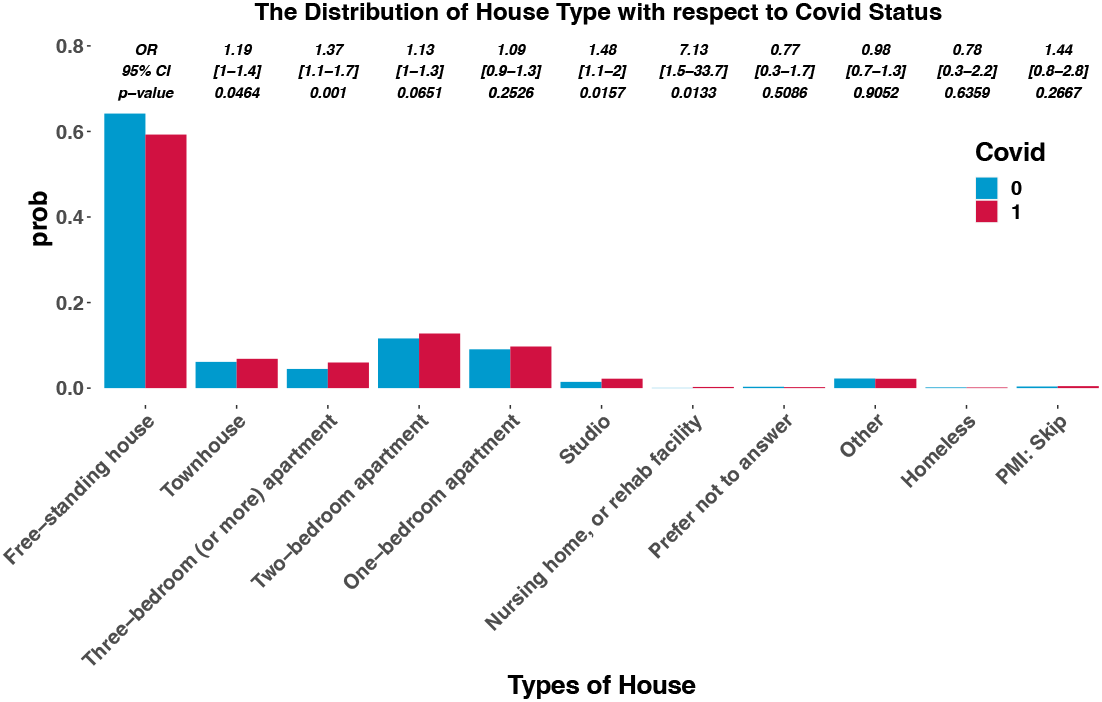
Distribution of house build type conditioned on COVID-19 status. A free-standing house has a less conditional probability of infection than non-infection, but the house types with shared components have a larger conditional probability of infection than non-infection. Odds ratios and p-values of COVID-19 infection for the house types compared to free-standing houses are shown above the probability bar graphs of the corresponding house type.

### 3.2 Associations between household type and mental health during COVID-19 pandemic

Next, we examined the impact of household type on mental health, specifically anxiety and stress. We hypothesized that COVID-19 status and infection risk may affect participants’ mental health by varying degrees; thus, the household type may affect a person’s mental health even though they are not infected, based on results from 3.1. Thus, we modeled the mental health outcome of anxiety or stress statuses using a multiple logistic regression model to evaluate how household types would affect the two conditions while controlling for various cofounders. The models included race, ethnicity, sex at birth, birth year, hygiene, social distancing behavior (e.g., shopping and outing behaviors), COVID-19 status, survey versions, plus the other mental health condition (e.g., stress to anxiety or vice versa) (See Methods). A large number of positive anxiety and stress cases were reported, consisting of 19,918 anxiety cases from 14,818 (23.6% of 62,664) participants and 21,821 stress cases from 15,810 (25.2% of 62,664) participants. We made a nonanxiety control cohort of 14,818 (896 partial matches) and a non-stress control cohort of 17,276 (933 partial matches) with matched race, sex at birth, age group, and survey versions (partial matches all maintained matched age groups).

The conditional probability of household type with respect to anxiety and stress is shown in Figs. 2 & 3. Household type affects anxiety and stress similarly to COVID-19 infection: participants living in a free-standing house comprised a smaller proportion among mental health cases as compared to mental health controls, while larger proportions of mental health cases were found in household types with a shared component. Participants who lived in the household type with a shared component showed an elevated risk of anxiety (OR=1.26, 95% CI 1.1 to 1.4; p=1.1×10^−6^) and stress (OR=1.29, 95% CI 1.2 to 1.4; p=4.3×10^−10^). For instance, participants in one-bedroom apartments (anxiety: OR = 1.48, 95% CI 1.3 to 1.7, p =2.4×10^−6^; stress: OR = 1.46, 95% CI 1.3 to 1.7; p=4.2×10^−8^) were found to have a significantly elevated risk of mental health conditions than participants who lived in free-standing houses; homeless had the highest odds ratio of anxiety (OR = 4.13, 95% CI 1.5 to 11.7; p = 0.008), particularly for those above their 50s. Race, sex, and age were confounding factors for the association between household type and mental health (e.g., stress) as all were significantly (p<0.01) associated with mental health in the matched models. COVID-19 status, household occupancy, hygiene, and social distancing habits (close contact, days at home and at work), were associated with both anxiety and stress (p<0.05). Last, stratification studies of sex and age group led to similar trends (data not shown), and anxiety and stress were correlated.

**Fig. 2.**
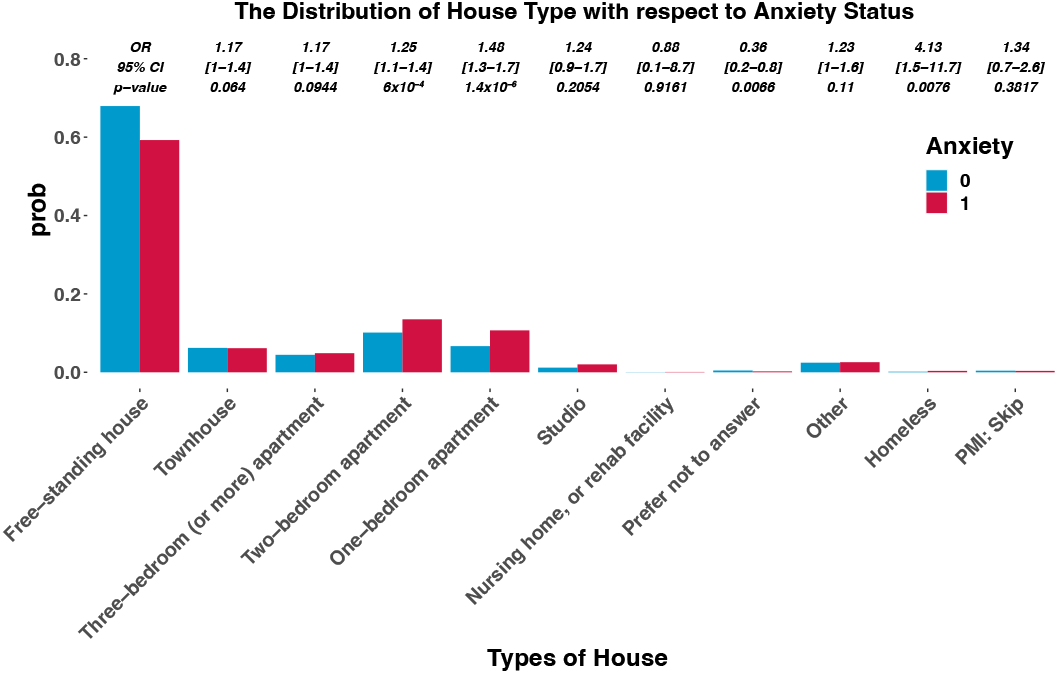
Distribution of house build type conditioned on anxiety status. Participants with anxiety are less likely to live in a free-standing house and more likely to live in a house type with shared components compared to those without anxiety. Odds ratios and p-values of anxiety for each of the house types as compared to free-standing houses are shown above the probability bar graphs for their corresponding type.

**Fig. 3.**
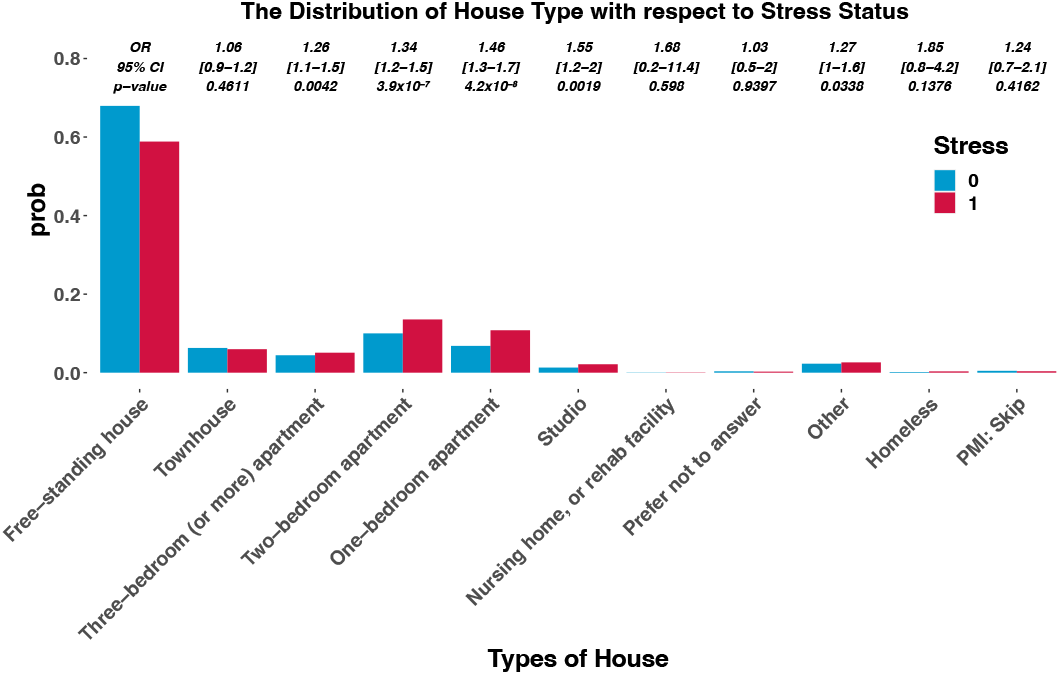
Distribution of house build type conditioned on stress status. Participants with stress are less likely to live in a free-standing house and more likely to live in a house type with shared components compared to those without stress. Odds ratios and p-values of stress for each of the house types as compared to free-standing houses are shown above the probability bar graphs for their corresponding type.

### 3.3 Impact of shopping behavior on COVID-19 status and mental health

Finally, we examined the association between COVID-19 status and nonessential shopping, or outings, using the same analysis in 3.1 & 3.2. Most shopping was done indoors; thus, it is an issue also related to the built environment. Fig. 4 shows the distribution of nonessential shopping and outings behavior with respect to COVID-19 status, which suggests more shopping and outings were associated with an elevated risk of COVID-19 infection. During the early stages of the pandemic, most participants tended to eliminate nonessential shopping. More than 70% of participants reported none of the days in the last five days, whether they got infected or not. Therefore, we set this shopping type as the reference level. The multiple logistic regression analysis showed that participants who went on outings more often had a higher risk of infection. Participants who shopped most days (more than three days within the last five days) yielded nearly 36% more risk as compared to the participants who shopped none of the days (OR = 1.36, 95% CI 1.1 to 1.8, p = 0.02). Interestingly, participants with frequent shopping behavior were not significantly associated with more anxiety or stress. The results suggest that commercially built environments, such as shopping malls, were likely to contribute to COVID-19 transmission in the initial stage of the pandemic. However, a more precise measurement of the variables is needed for a more robust conclusion.

**Fig. 4.**
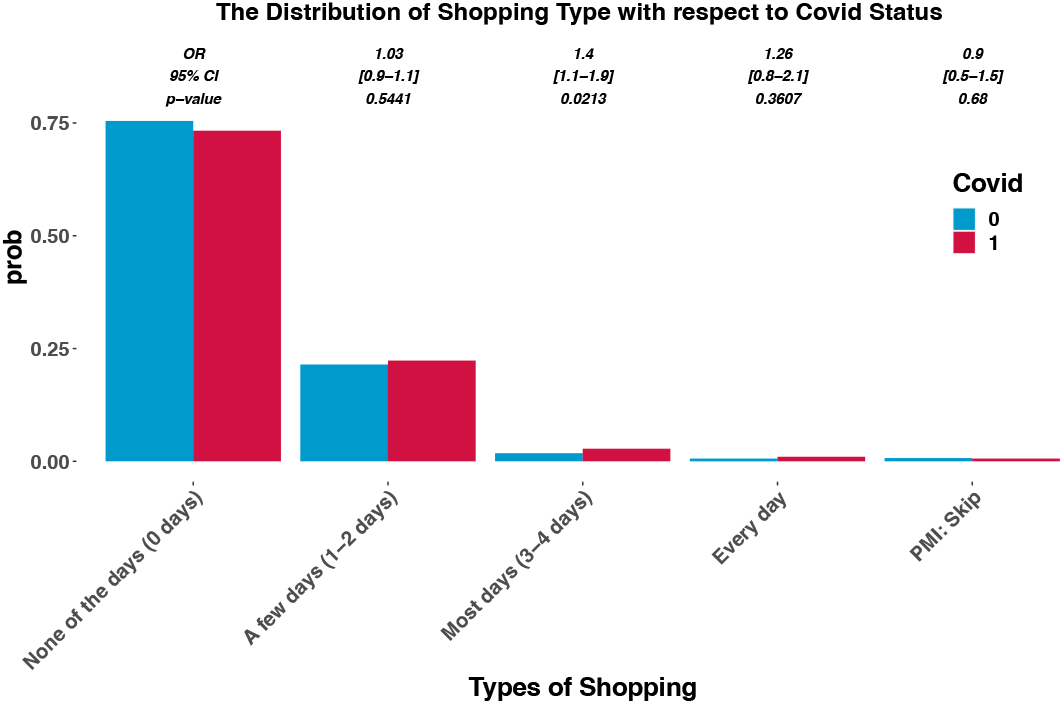
Distribution of just for fun shopping and outing behaviors in the last five days for the survey conditioned on COVID-19 status.

## 4 Discussion

In this study, we applied multiple logistic regression analyses using the COPE survey data and found that household type was associated with COVID-19 infection and mental health (e.g., anxiety) during the early stages of the COVID-19 pandemic. Our analysis also revealed that individuals with more frequent shopping or outings were more likely to get infected with COVID-19. Fig. 5. illustrates the summarized findings among the household type, COVID-19 infection, mental health, and nonessential shopping.

**Fig. 5.**
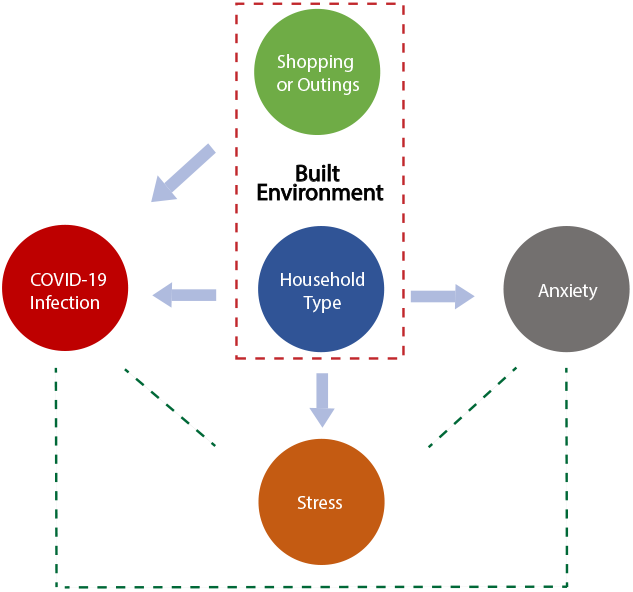
Relationships among the household type, COVID-19 infection, mental health, and non-essential shopping or outings. Arrows indicate associations between the built environment and its impact outcomes, while dotted lines show the confounding associations among the outcomes.

People who lived in nursing homes or rehab suggested a much higher possibility of becoming infected with COVID-19 compared to those who lived in free-standing houses. This was true even with stratification analysis using the same age group (although they lacked power). For instance, we found OR=3.96, 95% CI 0.7 to 23.2, p=0.13 for those over the age of 60. Reports suggested crowding is a risky factor for infection and mortality^26^, particularly in nursing homes. In addition, apartment dwellers (i.e., three-bedroom (or more) apartments) showed a higher tendency for contracting COVID-19, even when considering the confounding factor of the number of occupants in the house (data not shown). Meanwhile, two-bed or one-bedroom apartments showed inconsistent and underpowered results in some age groups, likely due to lower household occupancy. Although contributions from confounding factors that were not modeled cannot be excluded (e.g., interactions with visitors and workers), the possibility of influence from the built environment is high due to the stringent controls and reproducibility of the results in the stratified analyses. Concerning mental health, people who are homeless and participants who lived in apartments with shared living spaces were at a higher rate of reporting anxiety than inhabitants in free-standing homes. This corroborates prior reports and is partially caused by the congregate living conditions^27^. All the results indicated that built environments with shared components played a role in the transmission of SARS-CoV-2 and imposed mental burdens on dwellers during the COVID-19 pandemic.

Considering the association between social behaviors and COVID-19, such as outings and COVID-19 infection, there is an increasing trend of positive cases with more frequent nonessential shopping behaviors (3 or more days in the last five days) as compared to individuals who shopped for none of the days. As shown in Fig. 5, shopping and outing behaviors could relate to the built environment due to a large group of shoppers mixing indoors throughout the pandemic, providing common areas for COVID-19 viral transmission^28^.

In the early stages of the pandemic, increasing positive cases and deaths, a lack of knowledge of the virus, growing financial issues, strict social distancing regulations, and COVID-19 infections affected people’s well-being and daily life by contributing to widespread and increased psychological problems^20^. Further, previous studies have shown that people with pre-existing mental health conditions may have a higher tendency of being infected due to medical visits and emotional responses to the COVID-19 pandemic, thereby placing them at an increased risk of COVID-19 infection compared to those without mental health conditions^29^. Specifically, another study pointed out that individuals with mental health disorders are likely to have greater barriers in obtaining timely medical services, exacerbating mental health issues^30^. Our results corroborated previous findings (Fig. 5), although we could not distinguish the order (causal relationship) of mental disease and COVID-19 infection due to the lack of longitudinal information in the survey.

Although our study didn’t investigate them, latent connections also exist between shopping or outings with anxiety and stress. Previous studies have reported that moderate shopping could provide both psychological and therapeutic value^31,32^. Additionally, experiments about the effectiveness of diversion buying for stress release showed that a certain amount of spending was necessary to release stress^33^. Therefore, more just for fun shopping and outings may be related to anxiety and stress in a positive manner. Demonstrating this, frequent shoppers (every day) in their youth (20s age group) showed a reduction in stress (OR=0.07, p<0.0001).

The study is significant because it provides strong scientific evidence that demonstrates the associations between the built environment and COVID-19 transmission based on large-scale individualized data. While previous studies have suggested the associations based on summarized data (e.g., density of household types, COVID-19 transmission rate at the ZIP code scale^15^), the current study provided a more stringent investigation by controlling various confounders, such as age and sex.

The results of the study should be interpreted with caution due to various limitations: 1) The associations identified in the study may not indicate causal relationships, particularly from the secondary use of existing COPE data. 2) COVID-19 status was based on self-reporting in lieu of PCR-tested results, which were limited at the early stages of the pandemic. 3) COVID-19 infection is highly related to social behaviors, which vary significantly among age groups. Some of the stratification analyses in particular age groups were underpowered. 4) As shopping status is combined with outings, the impact on the commercial built environment is worth further study. 5) COVID-19, anxiety, and stress present interwoven relationships. Therefore, the order of the phenotypes can hardly be determined and are all confounded by socioeconomic status (e.g., employment status, financial difficulties, and other COVID-19 related impacts). This was not included in the current study due to the option for multiple answers rather than the multiple-choice survey in COPE, which might cause power issues and multicollinearity in regression.

In conclusion, the study demonstrated that the association of the built environment with a shared component tends to increase COVID-19 transmission and that their dwellers experience increased anxiety and stress levels. It is crucial to improve the quality of the built environment through planning, design, and management, pursuing a more resilient society that is able to cope with future pandemics.

## Data Availability

All data produced in the present study are available upon reasonable request to the authors.

https://www.researchallofus.org/data-tools/workbench/

## Authors’ Contributions

HL, BY, and SL conceived the study. HL and WL conducted the data processing and analysis. WL and HL drafted the manuscript. BY, EB, and AJ revised the manuscript. All authors reviewed the manuscript.

## Acknowledgements

This research has been conducted using All of Us Researcher Workbench platform The All of Us Research Program is supported by the National Institutes of Health, Office of the Director: Regional Medical Centers: 1 OT2 OD026549; 1 OT2 OD026554; 1 OT2 OD026557; 1 OT2 OD026556; 1 OT2 OD026550; 1 OT2 OD 026552; 1 OT2 OD026553; 1 OT2 OD026548; 1 OT2 OD026551; 1 OT2 OD026555; IAA #: AOD 16037; Federally Qualified Health Centers: HHSN 263201600085U; Data and Research Center: 5 U2C OD023196; Biobank: 1 U24 OD023121; The Participant Center: U24 OD023176; Participant Technology Systems Center: 1 U24 OD023163; Communications and Engagement: 3 OT2 OD023205; 3 OT2 OD023206; and Community Partners: 1 OT2 OD025277; 3 OT2 OD025315; 1 OT2 OD025337; 1 OT2 OD025276. In addition, the All of Us Research Program would not be possible without the partnership of its participants.

